# Drug Checking Services in Latin America vs. the United States: Staff Perspectives, Program Characteristics, and Barriers to Implementation

**DOI:** 10.64898/2026.04.11.26350657

**Authors:** John A. Bird, Joseph G. Rosen, Jaime Arredondo Sanchez Lira, Traci C. Green, Ju Nyeong Park

## Abstract

**Background:** Drug checking services (DCS) promote drug supply awareness among people who use drugs (PWUD) by detecting adulterants such as fentanyl and xylazine that are associated with overdose morbidity and mortality. However, there is limited research on DCS implementation in Latin America (LA).

**Methods:** We conducted a survey of 38 DCS across LA (n=10) and the US (n=28) and compared program characteristics and barriers between these two regions. We also conducted a focus group discussion (FGD) with staff representing six organizations implementing DCS in LA. FGD themes were mapped to constructs quantitatively assessed in the survey.

**Results:** Compared to US DCS, LA DCS more frequently reported funding gaps as a major implementation barrier (80% vs. 54%), law enforcement confiscating DCS supplies (38% vs. 11%), as well as offering supervised drug consumption (30% vs. 4%) and mental health/counseling (40% vs. 18%), but less frequently reported that DCS equipment was legal (44% vs. 75%). DCS on the Mexico-US border focused on people who inject drugs and offered syringe services, supervised consumption, and rapid sexually transmitted infection testing. DCS in central Mexico, Colombia, Peru, and Chile primarily provided DCS for the nightlife community (e.g., attendees of concerts/raves). Barriers to DCS implementation cited by FGD discussants included inadequate funding, DCS legal ambiguities, lack of government support, and cartel violence.

**Conclusion:** DCS in LA would benefit from increased funding, government support, and a more permissive legal environment, thereby strengthening harm reduction efforts and improving safety for PWUD.

## 1. Background

The overdose epidemic is a well-known public health crisis: while the number of fatal drug overdoses in the US has decreased from a peak of almost 108,000 in 2022 to just under 80,000 in 2024, they remain a top-ten leading cause of death (Garnett et al., 2026, Garnett et al., 2024, Xu et al., 2026). An estimated 62% of overdose deaths involve synthetic opioids such as fentanyl. The introduction of illicitly-manufactured fentanyls has also increased overdose risk for PWUD on the Mexico-US border (Friedman et al., 2022; Arredando et al., 2025; Gonzalez-Nieto et al., 2023).

Aggravating the fentanyl crisis are alpha-2-agonists such as xylazine, the veterinary sedative colloquially known as “tranq” or *“anastesia de caballo”* (horse anesthesia) in Spanish (Pacheco et al., 2025). The combination of fentanyl and xylazine, often present in the illicit drug supply as a comixture, has been referred to as a syndemic (Zhu et al., 2024). Because xylazine is a non opioid sedative, its effects cannot be reversed with naloxone (an opioid antagonist). Though xylazine initially emerged in opioid supplies in Puerto Rico in 2001, it later appeared in the mainland US’ opioid supply and is now also present along the Mexico-US border (Torruella, 2011, Zhu, 2023, Pacheco et al., 2025).

In recent years, Latin America’s nightlife scene has seen the arrival of “tusi,” also known as “tusibí” or “pink cocaine” (Fitzgerald et al., 2025). Tusi and tusibí are phonetic translations of 2C/2CB (Palamar, 2023). Contrary to its names, tusi generally contains neither 2C series drugs nor cocaine and is often a mixture of various substances, most commonly ketamine, MDMA, and caffeine (Fitzgerald et al., 2025; Díaz Moreno et al., 2022; Échele Cabeza, 2023). However, drug checkers have also found that “tusi” can contain methamphetamine, cocaine, opioids, and other substances (Palamar, 2023).

DCS were pioneered in the “party drug” scene in Europe in the 1990s and have the potential to enhance safety among PWUD in the context of an opaque illicit drug market (Laing, et al., 2018). These programs allow PWUD to have their drugs chemically analyzed for the presence of substances such as fentanyl, xylazine, and methamphetamine, facilitating more informed decisions about drug use. Drug checking (DC) staff typically pair analysis results with tailored harm reduction advice, and aggregated DC results enable public health surveillance of the drug supply (Maghsoudi, et al., 2021; Laing, et al., 2018).

In recent years, DCS have been used in response to the opioid overdose crisis (Laing, et al., 2018). One prior study showed that DC for fentanyl was associated with positive behavior change among women who inject drugs on the Mexico-US border: knowing that a sample contained fentanyl led half of the women in the study (30 total) to use less (14) or not use at all (1) (Goodman-Meza et al., 2022). Another study showed that among over 300 PWUD in eastern US cities, 90% felt that DCS would help them prevent an overdose (Sherman, et al., 2019).

This emerging body of evidence demonstrates the harm reduction benefits of DCS implementation and scale-up in LA and the US. However, in a 2021 systematic review of studies on DCS, only 1 of 90 included studies was conducted in LA (Maghsoudi et al., 2021). This suggests either a neglect of the region in research on DCS and/or a dearth of DCS programming in the region. Through this study, we sought to further characterize LA DCS, compare them to US DCS, and explore barriers to their implementation.

## 2. Methods

### 2.1. Survey

We conducted an online survey of DCS between November 2023 and January 2024, seeking participation from individuals representing discrete DCS. Participants were recruited from the Alliance for Collaborative Drug Checking, which originated in the US. This group defines itself as “a learning community for anyone working on initiatives or interested in expanding drug checking in their community regardless of the technologies that you are using” (Alliance for Collaborative Drug Checking, 2023). Eligible DCS had to be located in the Americas and be operating at the time of the survey. Diagnostic laboratories working exclusively with biological specimens and law enforcement agencies were ineligible. Staff self-completed the survey online in English, and we assisted Spanish-speaking DCS staff when necessary.

We used REDCap to administer the 20-30-minute survey, an earlier iteration of which was administered exclusively to North American programs in 2022 (Park et al., 2023). The survey’s questions spanned five domains: program location and attributes, operational characteristics, staffing and personnel, outcomes to date (e.g., total number of samples tested), and barriers and facilitators to implementation. Survey data was downloaded to Stata Version 17 (StataCorp LLC, College Station, Texas) for analysis. We created an LA group (n=10) and a US group (n=28) to compare frequencies and percentages of survey responses. Within the LA group, there were DCS from Mexico (n = 6), Colombia (n = 2), Peru (n = 1), and Chile (n = 1).

### 2.2. Focus Group Discussion

We moderated a 110-minute FGD in Spanish in September 2023 with DCS service providers and laboratory technicians in Mexico (n = 3), Colombia (n= 1), Peru (n = 1), and Chile (n = 1). The first author visited two DCS on the Mexico-US border in July 2023 as part of discussant recruitment. The remainder of service providers were sampled from the Alliance for Collaborative Drug Checking.

The FGD was recorded on Zoom and was transcribed automatically using Microsoft Transcribe. We later re-watched the FGD recording to verify the transcript’s accuracy. The quotes in the results section were translated to English using Google translate and edited for accuracy and clarity. We used Immersion-Crystallization, an inductive, iterative method to identify themes, categories, and patterns in the data (Borkan, 2022). These themes were mapped to the quantitative survey results, with a specific focus on barriers to implementation as well as program characteristics such as funding sources, target audiences, and other services offered by DCS.

### 2.3. Ethics

The study was approved by the Brown University Health Institutional Review Board. Survey participants received $25 USD and FGD discussants received $50 USD.

## 3. Results

### 3.1. Program types and drug checking specifics

Program characteristics varied widely by geography. Both FGD discussants located near the Mexico-US border worked at DCS that offered test strips for fentanyl, methamphetamine, and benzodiazepines; one offered them for xylazine as well. One of these DCS had a partnership with the Mexican government for mass spectrometry and gas chromatography.

The DCS in central Mexico, Colombia, Peru, and Chile primarily used colorimetry (i.e. color reagents), though the DCS in central Mexico and Colombia also had access to thin-layer chromatography on contract. At the time of the FGD, the DCS in Colombia had recently started using test strips for fentanyl and benzodiazepines as well.

US DCS more frequently reported they were stationary (46% vs. 0%) and offered on-site testing (89% vs. 60%) and off-site (i.e. laboratory) testing (64% vs. 20%). LA DCS more frequently reported they were mobile (50% vs. 7%) and offered self-testing, or “take-home,” kits (50% vs. 43%).

US DCS more frequently reported being part of a health department (14% vs 0%) or a university (11% vs. 0%), while LA DCS more frequently reported being community-based organizations (78% vs. 61%).

### 3.2. Populations served

The two FGD discussants working on the Mexico-US border focused primarily on serving people who inject drugs, mostly fentanyl and methamphetamine (the latter of which was sometimes smoked). Both sites stated that fentanyl has completely dominated the illicit opioid market in their respective geographies.

A substantial proportion of these programs’ clients were unhoused, often living in encampments. Many were either people who have been deported from the US, internal migrants, or cross-border migrants passing through Mexico en route to the US. Sex workers, including transgender women who sell sex, were also a key client population.

*“Most of the people we serve, I think 70-80%, live on the street… and they move in these contexts of sex work, substance use, [and] migration. Most of them are people who have been deported [from the US]”* - DCS staff, Mexico-US Border One of these DCS programs on the border stated that it served adults across a wide age range, from 17 to 70 years-old. The same DCS site on the border also served people from the US who do not have access to harm reduction services and crossed the border into Mexico to buy and use drugs more safely.

*“We also have a significant number of people who are from the US and who cross the border daily to buy and use substances [in Mexico]. They use the services of both the [supervised consumption] facility and the [harm reduction] supplies, since in this part of the US, there are no harm reduction programs.”* - DCS staff, Mexico-US Border

The FGD discussants in central Mexico, Colombia, Peru, and Chile provided services for the nightlife community (e.g., attendees of music festivals, concerts, and raves). Most often, these DCS offered their services at stands or booths at these events. The DCS in Chile estimated that most of their clients are between the ages of 20 and 40 years-old. The DCS in Peru stated that they most commonly check MDMA, cocaine, tusi, and less frequently, LSD. The vast majority (96%) of DCS surveyed in the US list people who use opioids, people who use stimulants, and people who deal/sell drugs as target audiences. In contrast, only 40% of LA DCS listed people who use opioids and people who deal/sell drugs as target audience; 80% of LA DCS listed people who use stimulants as their target audience.

The most common drug brought in to be tested at US DCS programs was fentanyl (79%), compared to 20% at DCS in LA. Other notable differences in common drugs were between crack cocaine (39% in the US vs. 0% in LA) and xylazine (25% in the US vs. 0% in LA).

### 3.3. Other services offered by DCS

In addition to DC, the two DCS near the Mexico-US border offered supervised drug consumption, syringe services programs, and rapid sexually transmitted infection testing (e.g. HIV and hepatitis). The larger of these two sites also offered medical care (with a physician and nursing staff), nutrition support, psychology/counseling services, and social services. This site also offered supervised consumption for both injection and inhalation drug use. The other, smaller site on the border offered supervised consumption, specifically for women who inject drugs.

The DCS in central Mexico, Colombia, Peru, and Chile offered information about safe substance use (e.g., dosing, expected effects, side effects) but not the types of robust services offered by the two DCS near the Mexico-US border.

Survey data showed that more US DCS than LA DCS offered syringe service programs (75% vs. 30%), housing/shelter (7% vs. 0%), medications for opioid use disorder (32% vs. 10%), naloxone (82% vs. 30%) rapid HIV testing (54% vs. 30%), rapid HCV testing (39% vs. 30%), HIV pre- and post-exposure prophylaxis (43% vs. 20%), case management (39% vs. 10%), recovery support coaching (28% vs. 10%), and laboratory DCS (25% vs. 10%). In contrast, LA DCS tended to offer supervised drug consumption (30% vs. 4%) and mental health/counseling (40% vs. 18%).

### 3.4. Barriers to program implementation

#### 3.4.1. Funding precarity

Most FGD discussants stated that funding gaps were a significant operational barrier. Consequently, they were understaffed and lacked advanced onsite DC technology (e.g. Fourier transform infrared spectroscopy [FTIR]), instead relying on test strips and colorimetry in their services.

*“More than 50% of our problems are related to money… For example, the lack of a physical space for us to conduct analyses and serve a larger and more diverse population at all times… Things like being able to dedicate more people to full-time work… Buying more expensive equipment, like an FTIR or something similar, seems impossible for us.”* - DCS staff, Chile

*“The budget challenge is also a significant factor. We would also like to have greater capacity to perform these analyses. As I mentioned, we don’t send samples in large quantities, but rather in small amounts, so we can’t do everything we’d like due to financial constraints.”* - DCS staff, Mexico-US border.

Our survey data shows that LA DCS cited funding as a major barrier with more frequency than DCS in the US (80% vs. 54%). Relatedly, LA DCS cited funding as a major facilitator with much less frequency (10% vs. 61%).

DCS in the US reported receiving funding from state/provincial health or other departments (36% vs. 0%) or local/regional health or other departments (25% vs. 0%); none of the LA DCS surveyed received such funding. LA DCS reported receiving funding from private philanthropy/foundations (40% vs. 21%) and obtaining payments from service users (20% vs. 11%).

*“Since 2018, the subsidies for harm reduction programs in Mexico have ended, and although there have been some small government interventions in harm reduction, these are truly insignificant compared to the need that exists in the country, right?”* - DCS staff, Mexico-US Border

### 3.5. Fear of violence

Both FGD discussants near the Mexico-US border cited fear of reprisal from organized criminal networks (i.e., cartels) as a barrier to service delivery. This was discussed as a challenge specifically during the beginning of the fentanyl crisis.

*“At first, the question was, how are we going to handle the issue of heroin being contaminated with fentanyl, for example? Our priority has always been to inform the community, but then we started to consider how we would do that without causing panic, because at first, people were saying, ‘Oh, I’m going to go to the dealer and tell him that what he’s selling me is contaminated.’”* - DCS staff, Mexico-US Border

The fear of violence was identified specifically as a barrier to DCS operation in the community, outside of the walls of the brick-and-mortar DCS site:

*“This violence, what we’re experiencing at the border, these conflicts over territory between different cartels… the truth is, we do feel vulnerable, and we feel like we’re at risk, especially if we go out and do this DC program on the streets or at other festivals. We’ll probably face some repercussions, like these groups finding out and coming here to do something to us.” -* DCS staff, Mexico-US Border

### 3.6. The rise of tusi

A FGD discussant from Colombia cited the rise of tusi as a challenge; the fact that it is a polydrug complicates both analysis and counseling.

*“Colorimetric tests are designed to determine a single substance. Here in Colombia, we have a very popular cocktail of substances called tusi, where it’s quite complicated to know what it might contain with the reagents because the analysis is still preliminary. With MDMA crystals, the result should match the person’s expectations, but with this cocktail of substances, it’s much more difficult because we first have to educate people about what it is, and given that [it’s a cocktail], we can’t even give a positive or negative result.”* - DCS staff, Colombia

### 3.7. Lack of government support, criminalization, and legal uncertainty

Multiple discussants lamented the government’s criminalization of substance use and lack of support for harm reduction as a public health strategy.

*“The agency in charge of drugs doesn’t belong to the Ministry of Health, it belongs to the Ministry of the Interior [similar to the US Department of Homeland Security]… almost like the defense of the country. That’s how the government views it, not like a health problem but like an invasion.” - DCS staff, Chile*

*“The criminalization of substance use, and of PWUD, makes it very difficult for them to access healthcare. They will be stigmatized and criminalized in those spaces. And as for our national policy? The President of the Republic, speaking about it here in Mexico, doesn’t value PWUD. He [former president López Obrador] criminalizes drug use as something very bad. And it doesn’t seem like there will be any real possibility of including harm reduction as a national policy… Before, at least there were resources that could be allocated to organizations. Now the government doesn’t pay a single peso for a single syringe… Those resources come from abroad, or from other institutions.”* - DCS staff, Mexico-US border

Uncertainty due to change in governments was another factor cited, both in Chile and Colombia.

*“It’s like we have to juggle things, and that’s how we have to fit in with the political stance of the government in power at any given time, for as long as it lasts, and because of the short-lived governments that don’t have time to implement much. So, even if a government wants to facilitate our access to this equipment, at any moment it ends, another one comes in, and that’s where all the progress is left.”* - DCS staff, Chile

*“For example, let’s say with [the decriminalization of] cannabis, we were kind of taking a step forward. But it’s going to get complicated, right? … People might make decisions that could be lost when the right-wing returns to power, like many of the things we have now, and that’s part of our fear.”* - DCS Staff, Colombia

Another barrier cited, specifically in Peru and Chile, was uncertainty regarding the legality of DC supplies, of drugs themselves, and of paraphernalia.

*“Drug use isn’t penalized in Chile, but you can’t possess it – you can consume it, but you can’t have it, so… how does someone get from their home to the DCS without breaking the law?”* - DCS staff, Chile

*“There is a certain ambiguity in the work we do; it’s not illegal, but it’s not exactly legal either.”* - DCS staff, Peru

Our survey showed that LA DCS more frequently reported having had DC supplies confiscated by law enforcement (38% vs. 11%) and to report that DC equipment was neither legal nor illegal (44% vs. 7%). LA DCS reported that DC supplies were legal in their state/territory with less frequency than US DCS (44% vs. 75%).

### 3.8. An ideal world for drug checking

When asked about what an ideal world would be in relation to DCS, FGD discussants agreed that an ideal world would be one in which their work was not needed, and one in which governments supported their work instead of impeding it. Specifically, they cited increased funding, decriminalization of drug use, and having a state-regulated drug supply as ways in which governments could promote their work.

*“The ideal world would be one where public policy [around DCS] is closely linked to health services, which is what we’re most hopeful for. And let’s say that the most radical solution is that there would be a safe drug supply through which people can access their substances legally with state oversight.”* - DCS staff, Colombia

*“I share my colleagues’ opinion of not having to do this work… but if we do have to do it, we would like to have an environment free of criminalization for drug use, so that people can easily access our services without fear of being extorted by the police for carrying just one syringe, or for carrying the minimum dose for consumption.”* - DCS staff, Mexico-US border

## 4. Discussion

As opposed to their US counterparts, LA DCS tended not to offer onsite-testing and laboratory testing; they were also less often operating in a stationary setting or associated with a public health department or university. DCS in LA more frequently endorsed funding gaps as a major barrier to operation compared to the US. A lower proportion of LA DCS received funding from state/provincial health or local/regional health departments compared to their counterparts in the US. Our FGD discussants in LA agreed that funding precarity significantly impaired their ability to adequately staff their organizations and purchase desired DC technologies (e.g. FTIR). Increased funding would allow DCS in LA to obtain more advanced DC technologies (e.g. FTIR) and facilitate access to laboratory-based DC. This could facilitate the generation of data to improve comparison between DCS as well as public health surveillance of the drug supply to better characterize overdose risk.

With a few notable exceptions (namely supervised consumption and mental health/therapy), almost all additional harm reduction services were offered with less frequency by LA DCS compared to the US. These discrepancies are likely multifactorial: LA DCS reported both a lack of adequate funding and a dearth of institutional support, though they also served patient populations with different drug use patterns than US DCS. Because fentanyl is ubiquitous in the US illicit opioid supply, it is unsurprising that US DCS more frequently reported providing services tailored to people who use fentanyl: syringe services (fentanyl is commonly injected), STI testing (blood-borne STIs can be transmitted via injection), medications for opioid use disorder, and naloxone distribution for overdose reversal.

However, LA DCS more frequently reported offering supervised drug consumption. This may be a benefit of the ambiguity regarding harm reduction’s legality in the region. But legal ambiguity can also be detrimental: LA DCS more frequently reported confiscation of equipment and drugs/paraphernalia by law enforcement, and more frequently reported they were unsure whether their work was legal.

Within LA, the DCS on the border serve a marginalized population vulnerable to structural violence (Dubal, et al. 2021). As a result, additional services offered by border DCS are more robust and include medical care and social services. Because the DCS in central Mexico, Colombia, Peru, and Chile are focused on providing DCS for the nightlife community, they offered fewer additional services.

FGD discussants described an ideal world as one in which government policies supported, or even provided DCS, and in which they were linked to health services and clients could access services free of criminalization and stigmatization. Most discussants agreed they hoped for a world in which DCS were unnecessary, with two discussants articulating visions of a state-regulated drug supply.

### 4.1. Limitations

LA is an expansive, heterogeneous region, and it is impossible to fully characterize DCS across the region. Our survey participants and FGD discussants were drawn from only four countries: Mexico, Colombia, Peru, and Chile. We acknowledge the fraught task of comparing the entire region of LA to one country, the US. Another limitation is our small sample size, having conducted only one FGD and having only 10 participants from LA in our survey. Survey participants and FGD discussants were sampled from the same group, the Alliance for Collaborative Drug Checking. As such, we did not capture non-members. However, to our knowledge, this is the only survey of DCS in LA conducted since 2017 (Barrat et al., 2017). Our hope is that an expanded survey can be repeated in future years to document the important harm reduction work being carried out by these organizations.

### 4.2. Conclusions

LA DCS are operating in an extremely difficult environment characterized by barriers including inadequate funding, government opposition (or at best, indifference), and in some cases, fear of violence from organized crime networks (the latter specifically near the Mexico US border).

They would benefit from government support in the form of increased funding and decriminalization of DC equipment, drugs, and paraphernalia. These reforms would allow these programs to pay staff adequately, provide more technologically advanced DC, and offer additional wrap-around services to their clients, improving the health of PWUD. Further, a state-regulated drug supply could eliminate the incentives for an opaque illicit drug supply and the need for DCS.

**Table 1.** DCS offered, program descriptions, stationary vs. mobile (LA vs. US)

|  | Latin America |  | United States |  |
| --- | --- | --- | --- | --- |
| **What drug checking services are currently offered to clients through your program?** | **n=10** | **Percent** |  | **Percent** |
| Self-testing kits ("take home kits") | 5 | 50% | 12 | 43% |
| On-site testing | 6 | 60% | 25 | 89% |
| Off-site testing | 2 | 20% | 18 | 64% |
| Other | 1 | 10% | 5 | 18% |
| **Program description** | **n=9** |  | **n=28** |  |
| Health department | 0 | 0% | 4 | 14% |
| University | 0 | 0% | 3 | 11% |
| Community-based organization | 7 | 78% | 17 | 61% |
| Private lab | 0 | 0% | 1 | 4% |
| Other | 2 | 22% | 3 | 11% |

| Is your program stationary or mobile? | n=8 |  | n=28 |  |
| --- | --- | --- | --- | --- |
| Stationary | 0 | 0% | 13 | 46% |
| Mobile (i.e. Outreach van or portable equipment) | 4 | 50% | 2 | 7% |
| Both | 4 | 50% | 12 | 43% |
| Other | 0 | 0% | 1 | 4% |

**Table 2.** Target audience and drugs commonly tested (LA vs. US)

Target audience and drugs commonly tested (LA vs. US)
|  | Latin America |  | United States |  |
| --- | --- | --- | --- | --- |
| Who is the target audience of the program? | n=10 | Percent |  | Percent |
| People who use opioids | 4 | 40% | 27 | 96% |
| People who use stimulants | 8 | 80% | 27 | 96% |
| People who use psychedelics | 4 | 40% | 23 | 82% |
| People who use synthetic marijuana | 0 | 0% | 11 | 39% |
| Providers/first responders who care for PWUD | 2 | 20% | 8 | 29% |
| People who deal/sell drugs | 4 | 40% | 27 | 96% |
| Other | 1 | 10% | 2 | 7% |
| **What drugs are most commonly brought in to be tested?** | **n=10** | **Percent** |  | **Percent** |
| Fentanyl | 2 | 20% | 22 | 79% |
| Heroin | 3 | 30% | 12 | 43% |
| Cocaine (powder) | 4 | 40% | 19 | 68% |
| Crack cocaine | 0 | 0% | 11 | 39% |
| Methamphetamine | 4 | 40% | 18 | 64% |
| Xylazine | 0 | 0% | 7 | 25% |
| Pills | 5 | 50% | 16 | 57% |
| Other | 4 | 40% | 7 | 25% |

**Table 3:** Other services offered by DCS (LA vs. US)

|  | Latin America |  | United States |  |
| --- | --- | --- | --- | --- |
| **In addition to drug checking, are there other services that your program offers on site?** | **n=10** | **Percent** | **n=28** | **Percent** |
| Syringe services/exchange | 3 | 30% | 21 | 75% |
| Naloxone distribution | 3 | 30% | 23 | 82% |
| Supervised drug consumption | 3 | 30% | 1 | 4% |
| Rapid HIV testing | 3 | 30% | 15 | 54% |
| HIV PrEP/PEP | 2 | 20% | 12 | 43% |
| Rapid STI testing | 2 | 20% | 6 | 21% |
| Rapid HCV testing | 3 | 30% | 11 | 39% |
| Methadone/buprenorphine/naltrexone/MOU | 1 | 10% | 9 | 32% |
| Mental health/Counseling | 4 | 40% | 5 | 18% |
| Other medical services | 2 | 20% | 10 | 36% |
| Recovery support groups | 0 | 0% | 5 | 18% |
| Recovery support coaching | 1 | 10% | 9 | 32% |
| Housing/shelter | 0 | 0% | 2 | 7% |
| Case management | 1 | 10% | 11 | 39% |
| Laboratory testing services | 1 | 10% | 7 | 25% |
| Other services | 6 | 60% | 7 | 25% |

**Table 4:**
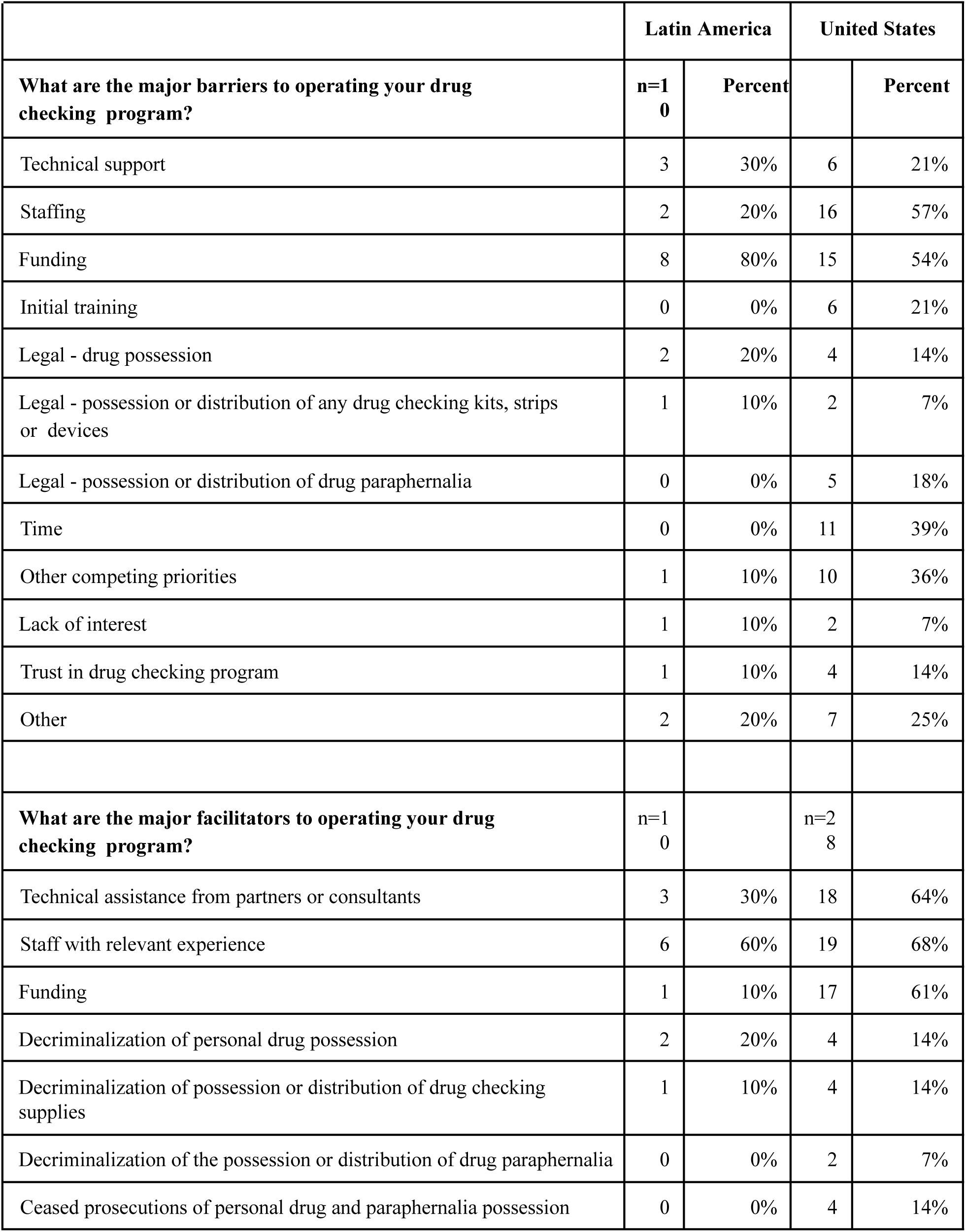
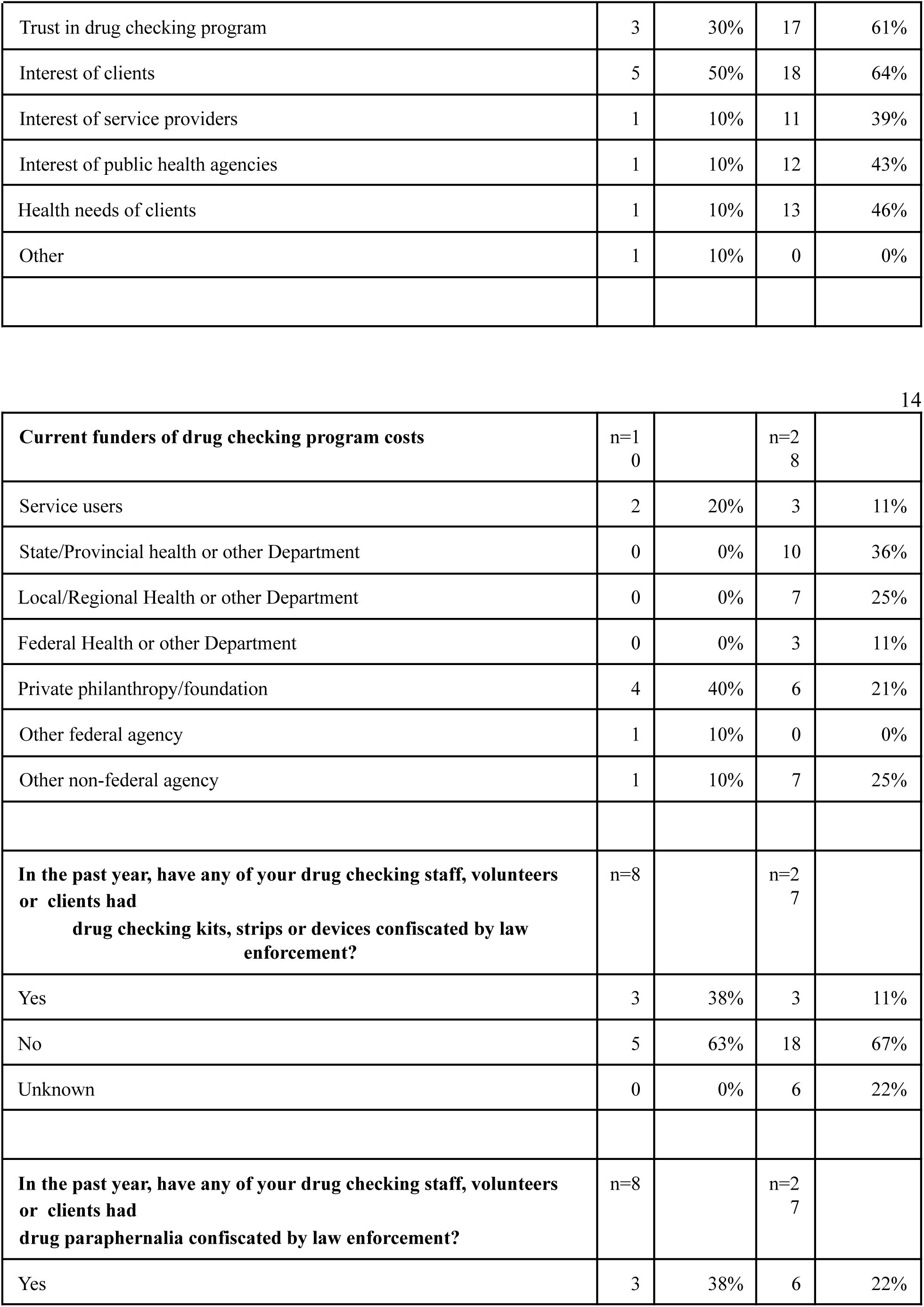
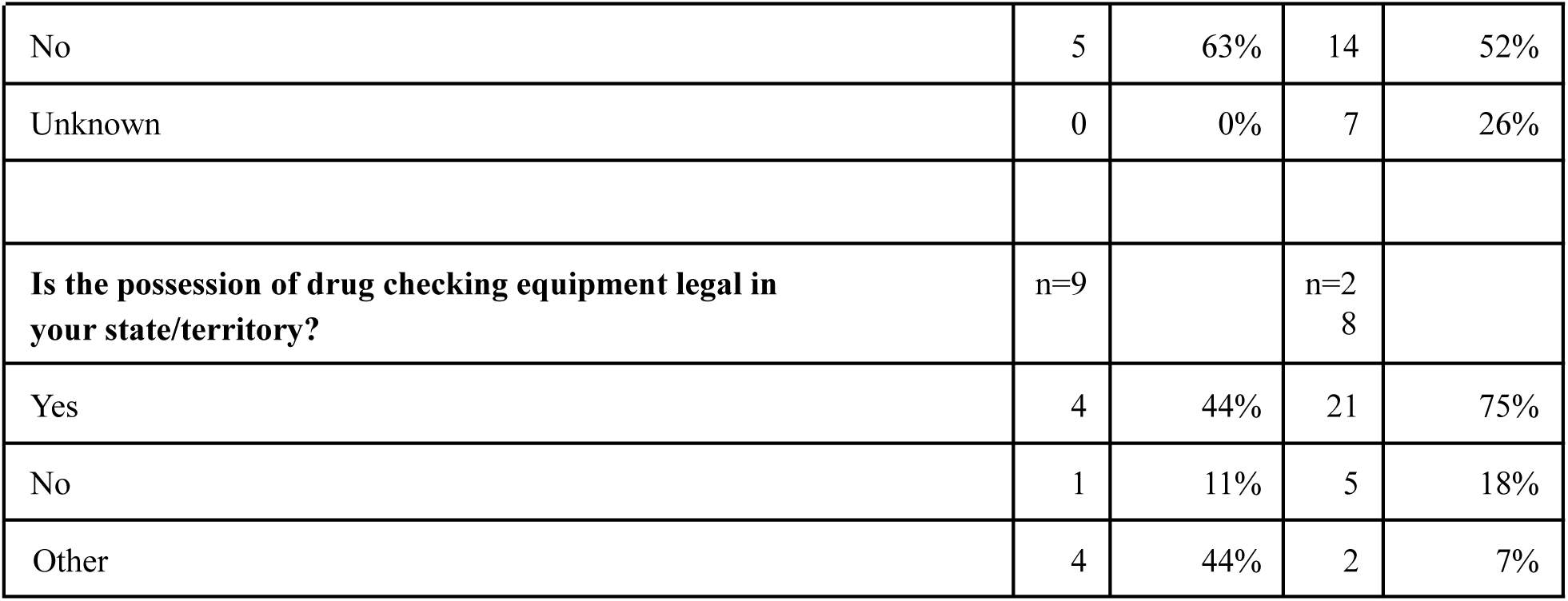
Barriers, facilitators, funders, confiscation by authorities, and drug checking legality (LA vs. US)

## Data Availability

All data produced in the present study are available upon reasonable request to the authors.

## Acknowledgement and funding

We are grateful to our survey participants and FGD discussants and to their clients. We thank Erin Thompson and Jessica Tardif for their contributions to data collection and project administration. JGR, TCG, and JNP were funded by the Center of Biomedical Research Excellence (COBRE) on Opioids and Overdose at Rhode Island Hospital, a program of the National Institute of General Medical Sciences (P20GM125507). JGR acknowledges funding from the National Institute of Mental Health (R25MH083620). JASL was funded by the Canada Research Chair Program. JAB was funded by Warren Alpert Medical School’s Primary Care Population Medicine Program.

## References

1. Arredondo Sanchez Lira, J., Chavez, A., Pacheco, L., 2025. The emergence of fentanyl in Tijuana: Implementing new public health interventions, in: Cutrona, S., Rossen, J. (Eds.), Fentanyl: From the Labs to the Streets. Routledge.

2. Bailey, K., Abramovitz, D., Artamonova, I., Davidson, P., Stamos-Buesig, T., Vera, C.F., Patterson, T.L., Arredondo, J., Kattan, J., Bergmann, L., Thihalolipavan, S., Strathdee, S.A., Borquez, A., 2023. Drug checking in the fentanyl era: Utilization and interest among people who inject drugs in San Diego, California. Int. J. Drug Policy 118, 104086. 10.1016/j.drugpo.2023.104086

3. Barratt, M.J., Kowalski, M., Maier, L.J., Ritter, A., 2018. Global review of drug checking services operating in 2017. National Drug and Alcohol Research Centre Bulletin No. 24. UNSW Sydney, Sydney.

4. Borkan, J., 2022. Immersion–crystallization: A valuable analytic tool for healthcare research. Fam. Pract. 39, 785–789. 10.1093/fampra/cmab158

5. Díaz Moreno, M., et al., 2022. Échele Cabeza as a harm reduction project and activist movement in Colombia. Drugs Habits Soc. Policy 23, 263–276.

6. Dubal, S., 2021. Policing, poverty, and punishment in the fentanyl era. Soc. Sci. Med. 289, 114405. 10.1016/j.socscimed.2021.114405

7. Échele Cabeza, 2023. Ingredientes del tusi y adulterantes en MDMA: Informe de resultados de análisis confirmativo GC/MS.

8. Fitzgerald, N.D., Abukahok, N., Palamar, J.J., 2025. When pink powders shift the drug landscape: tusi (“pink cocaine”) and other colored powders. Int. J. Drug Policy 146, 105044. 10.1016/j.drugpo.2025.105044

9. Friedman, J., Bourgois, P., Godvin, M., Chavez, A., Pacheco, L., Segovia, L.A., Beletsky, L., Arredondo, J., 2022. The introduction of fentanyl on the US–Mexico border: An ethnographic account triangulated with drug checking data from Tijuana. Int. J. Drug Policy 104, 103678. 10.1016/j.drugpo.2022.103678

10. Garnett, M.F., Miniño, A.M., 2024. Drug overdose deaths in the United States, 2003–2023. NCHS Data Brief No. 522. National Center for Health Statistics.

11. Garnett, M.F., Miniño, A.M., 2026. Drug overdose deaths in the United States, 2023–2024. NCHS Data Brief No. 549. National Center for Health Statistics.

12. Gonzalez-Nieto, P., et al., 2023. Intersections between syndemic conditions and stages along the continuum of overdose risk among women who inject drugs in Mexicali, Mexico. Harm Reduct. J. 20, 79.

13. Goodman-Meza, D., Arredondo, J., Slim, S., Angulo, L., Gonzalez-Nieto, P., Loera, A., Shoptaw, S., Cambou, M.C., Pitpitan, E.V., 2022. Behavior change after fentanyl testing at a safe consumption space for women in Northern Mexico: A pilot study. Int. J. Drug Policy 106, 103745. 10.1016/j.drugpo.2022.103745

14. Laing, M.K., Tupper, K.W., Fairbairn, N., 2018. Drug checking as a potential strategic overdose response in the fentanyl era. Int. J. Drug Policy 62, 59–66. 10.1016/j.drugpo.2018.10.001

15. Maghsoudi, N., Tanguay, J., Scarfone, K., Rammohan, I., Ziegler, C., Werb, D., Scheim, A.I., 2021. Drug checking services for people who use drugs: A systematic review. Addiction 117, 532–544. 10.1111/add.15734

16. Pacheco, L., Friedman, J., Bourgois, P., Chavez, A., Segovia, L.A., Beletsky, L., Arredondo, J., 2025. Xylazine and the changing landscape of the illicit drug supply: Implications for harm reduction. Harm Reduct. J. 22, 34. 10.1186/s12954-024-01143-2

17. Palamar, J.J., 2023. Tusi: A new ketamine concoction complicating the drug landscape. Am. J. Drug Alcohol Abuse 49, 546–550. 10.1080/00952990.2023.2207716

18. Park, J.N., Tardif, J., Thompson, E., Rosen, J.G., Arredondo Sanchez Lira, J., Green, T.C., 2023. A survey of North American drug checking services operating in 2022. Int. J. Drug Policy 121, 104206. 10.1016/j.drugpo.2023.104206

19. Sherman, S.G., Park, J.N., Glick, J.L., McKenzie, M., Morales, K.B., Christensen, T., Green, T.C., 2019. Acceptability of implementing community-based drug checking services for people who use drugs in three United States cities: Baltimore, Boston and Providence. Int. J. Drug Policy 68, 110–116. 10.1016/j.drugpo.2019.03.019

20. Torruella, R.A., 2011. Xylazine (veterinary sedative) use in Puerto Rico. Subst. Abuse Treat. Prev. Policy 6, 7.

21. Xu, J.Q., Murphy, S.L., Kochanek, K.D., Arias, E., 2026. Mortality in the United States, 2024. NCHS Data Brief No. 548. National Center for Health Statistics.

